# Analysis of B-cell receptor repertoire to evaluate the immunogenicity of SARS-CoV-2 RBD mRNA vaccine: MAFB-7256a (DS-5670d)

**DOI:** 10.1101/2024.07.19.24310697

**Authors:** Goh Ohji, Yohei Funakoshi, Kimikazu Yakushijin, Takaji Matsutani, Tomoki Sasaki, Takahiro Kusakabe, Sakuya Matsumoto, Taiji Koyama, Yoshiaki Nagatani, Keiji Kurata, Shiro Kimbara, Naomi Kiyota, Hironobu Minami

**Author notes:** **Corresponding author**: Yohei Funakoshi, M.D., Ph.D. Division of Medical Oncology/Hematology, Department of Medicine, Kobe University Hospital and Graduate School of Medicine. G.O., Y.F. and K.Y contributed equally to this work.

## Abstract

A monovalent Omicron XBB.1.5 mRNA RBD analogue vaccine, MAFB-7256a (DS-5670d), was newly developed and approved in Japan in the Spring of 2024 for preventing COVID-19. However, clinical efficacy data for this vaccine are currently lacking. We previously established the Quantification of Antigen-specific Antibody Sequence (QASAS) method to assess the response to SARS-CoV-2 vaccination at the mRNA level using B-cell receptor (BCR) repertoire assays and the Coronavirus Antibody Database (CoV-AbDab). Here, we used this method to evaluate the immunogenicity of MAFB-7256a. We analyzed repeated blood samples using the QASAS method from three healthy volunteers before and after MAFB-7256a vaccination. BCR response increased rapidly one week post-vaccination and then decreased, as with conventional vaccine. Notably, the matched sequences after MAFB-7256a vaccination specifically bound to the receptor-binding domain (RBD), with no sequences binding to other epitopes. These results validate that MAFB-7256a is an effective vaccine that exclusively induces antibodies specific for the RBD, demonstrating its targeted immunogenic effect.

## INTRODUCTION

In August 2021 and January 2022, the U.S. Food and Drug Administration (FDA) granted approval to mRNA vaccines targeting severe acute respiratory syndrome coronavirus 2 (SARS-CoV-2), namely Pfizer-BioNTech BNT162b2 and Moderna mRNA-1273. It was demonstrated that these mRNA vaccines, designed to elicit immune responses against the spike protein, were highly effective in preventing symptomatic Coronavirus Disease 2019 (COVID-19) during the initial phases of the pandemic.^1–3^ The spike protein of SARS-CoV-2, essential for receptor recognition and cell membrane fusion, comprises two subunits, S1 and S2.^4,5^ Both BNT162b2 and mRNA-1273 encode the full-length spike protein of SARS-CoV-2.^1–3^

The S1 subunit, consisting of 672 amino acids (residues 14-1273), includes an N-terminal domain (NTD) and the receptor-binding domain (RBD).^6,7^ The RBD, which is particularly crucial, facilitates virus-host cell interaction through its receptor-binding motif, which binds with the host cell’s angiotensin-converting enzyme 2 (ACE2).^6,7^ Another COVID-19 vaccine, a monovalent Omicron XBB.1.5 mRNA RBD analogue vaccine named MAFB-7256a (DS-5670d), has been developed by Daiichi Sankyo in Japan, and was approved by Japanese government in November 2023. The clinical and biological efficacy of this vaccine has not been fully evaluated, however, and further studies are required.

Previously, we established a novel method to assess the response to SARS-CoV-2 vaccination at the mRNA level by quantifying antigen-specific antibody sequences, named the “Quantification of Antigen-specific Antibody Sequence (QASAS) method”.^8,9^ This method uses B-cell receptor (BCR) repertoire data on activation of humoral immunity and a database of BCR sequences that bind to target antigens, such as the Coronavirus Antibody Database (CoV-AbDab).^10^ Since CoV-AbDab also contains information on strains and epitopes to which each antibody sequence binds, our method allows us to infer the strain and epitope specificity of antibody sequences generated by the vaccine.

Here, we used the QASAS method to evaluate immunogenicity after MAFB-7256a vaccination.

## MATERIAL AND METHODS

### Participants

Immune response following monovalent Omicron XBB.1.5 mRNA RBD analogue vaccination (MAFB-7256a [DS-5670d] [Daiichi Sankyo]) was evaluated by ELISA in 13 healthy volunteers (HV) enrolled in March-April 2024. All participants had a previous history of receipt of other COVID-19 vaccines. Among them, immunogenicity was tested in more detail using the QASAS method in three volunteers (MAFB-7256a HV 1-3). In addition, as a control, 14 participants with antigen exposure to SARS-CoV-2 (3 cases, SARS-CoV-2 infection; 9 cases, BNT162b2 vaccine, which encodes SARS-CoV-2 full-length spike protein; 2 cases, mRNA-1273 vaccines, which encodes SARS-CoV-2 full-length spike protein) were analyzed by the QASAS method (Supplementary Table 1).

The study protocol was approved by Kobe University Hospital Ethics Committee (No. B2356701). The study was conducted in accordance with the principles of the Declaration of Helsinki. The participants provided written informed consent for this research.

### Sample collection and processing

Serum samples were obtained by centrifuging blood samples for 10 min at 1000 × g at room temperature, and immediately transferred to a freezer kept at −80 °C.

Peripheral blood mononuclear cells (PBMCs) were isolated from peripheral blood samples using Ficoll-Paque Plus (GE Healthcare, Little Chalfont, UK) and stored at -80°C using CELLBANKER (Zenogen Pharma, Fukushima, Japan) until analysis. Total RNA was extracted with TRIzol LS (Thermo Fisher Scientific, Waltham, MA, USA) from PBMC for BCR repertoire analysis and purified with an RNeasy Mini Kit (Qiagen, Hilden, Germany) in accordance with the manufacturer’s instructions. RNA amounts and purity were measured with an Agilent 2200 TapeStation (Agilent Technologies, Santa Clara, CA, USA).

### Measurement of SARS-CoV-2 strain-specific IgG antibody titers

SARS-CoV-2 strain-specific anti-spike (anti-S) IgG titers were measured with an in-house ELISA using trimeric full-length spike proteins generated by a silkworm-baculovirus expression system, as previously described.^11^ Briefly, purified target proteins of the WT and Omicron (BA.1) strains were used as coating antigens for ELISA. Optical density at 450 nm (OD 450 nm) and 570 nm (OD 570 nm) was read using a microplate reader after ELISA, and the anti-S IgG titer of each sample was reported as the OD value (OD 450 nm − 570 nm).B-cell receptor repertoire analysis

BCR repertoire analysis was performed using unbiased next-generation sequencing developed by Repertoire Genesis, Inc. (Osaka, Japan).^12^ Briefly, cDNA was synthesized from total RNA using the polyT18 primer (BSL-18E) and Superscript III reverse transcriptase (Invitrogen, Carlsbad, CA, USA). After synthesizing double-strand (ds)-cDNA, the P10EA/P20EA dsDNA adaptor was ligated and cut with the *Not*I restriction enzyme. Nested PCR was performed with KAPA HiFi DNA Polymerase (Kapa Biosystems, Woburn, MA, USA) using IgG constant region-specific primers (CG1 and CG2) and P20EA. The amplicon library was prepared by amplification of the second PCR products using P22EA-ST1 and CG-ST1-R. Index (barcode) sequences were added by amplification with a Nextera XT Index Kit v2 Set A (Illumina, San Diego, CA, USA). Sequencing was performed using the Illumina MiSeq paired-end platform (2×300 bp). BCR sequences were assigned based on identity with reference sequences from the international ImMunoGeneTics information system® (IMGT) database (http://www.imgt.org) using repertoire analysis software originally developed by Repertoire Genesis, Inc. (Osaka, Japan).

### Database and SARS-CoV-2-specific sequence search

BCR/antibody sequences specific for SARS-CoV-2 were downloaded from CoV-AbDab (http://opig.stats.ox.ac.uk/webapps/covabdab/). Data updated on 13 June 2023, containing 12,536 entries, were used as reference. Antibodies with immunoglobulin heavy chain sequences with identical V and J genes and CDR3 amino acid sequences in the CoV-AbDab database were grouped as unique reference sequences. These unique reference sequences were classified based on their binding specificity to viruses, strains, and epitopes. Sequence classification was performed by matching virus search string (SARS-CoV-2), strain search strings (WT, Alpha, Beta, Gamma, Delta, Epsilon, Iota, Kappa, Lambda, Mu, Omega, Omicron) or epitope search strings (Supplemental Table 2) against the database registration information. Sequences detected in BCR repertoire analysis of PBMCs from vaccinated patients were searched against the classified reference sequence, and sequences with a CDR3 amino acid sequence exact match (Levenshtein distance 0, LV0) or 1 or 2 amino acid mismatch (LV1, 2) were counted.^8^

## RESULTS

### Serological outcomes after MAFB-7256a on anti-SARS-CoV-2 strain-specific spike IgG antibody measurement by ELISA

Anti-spike IgG antibody titers for original and Omicron strain were measured pre- and 14 days after MAFB-7256a vaccination in all participants, and titers were clearly increased after vaccination (Figure 1).

**Figure 1.**
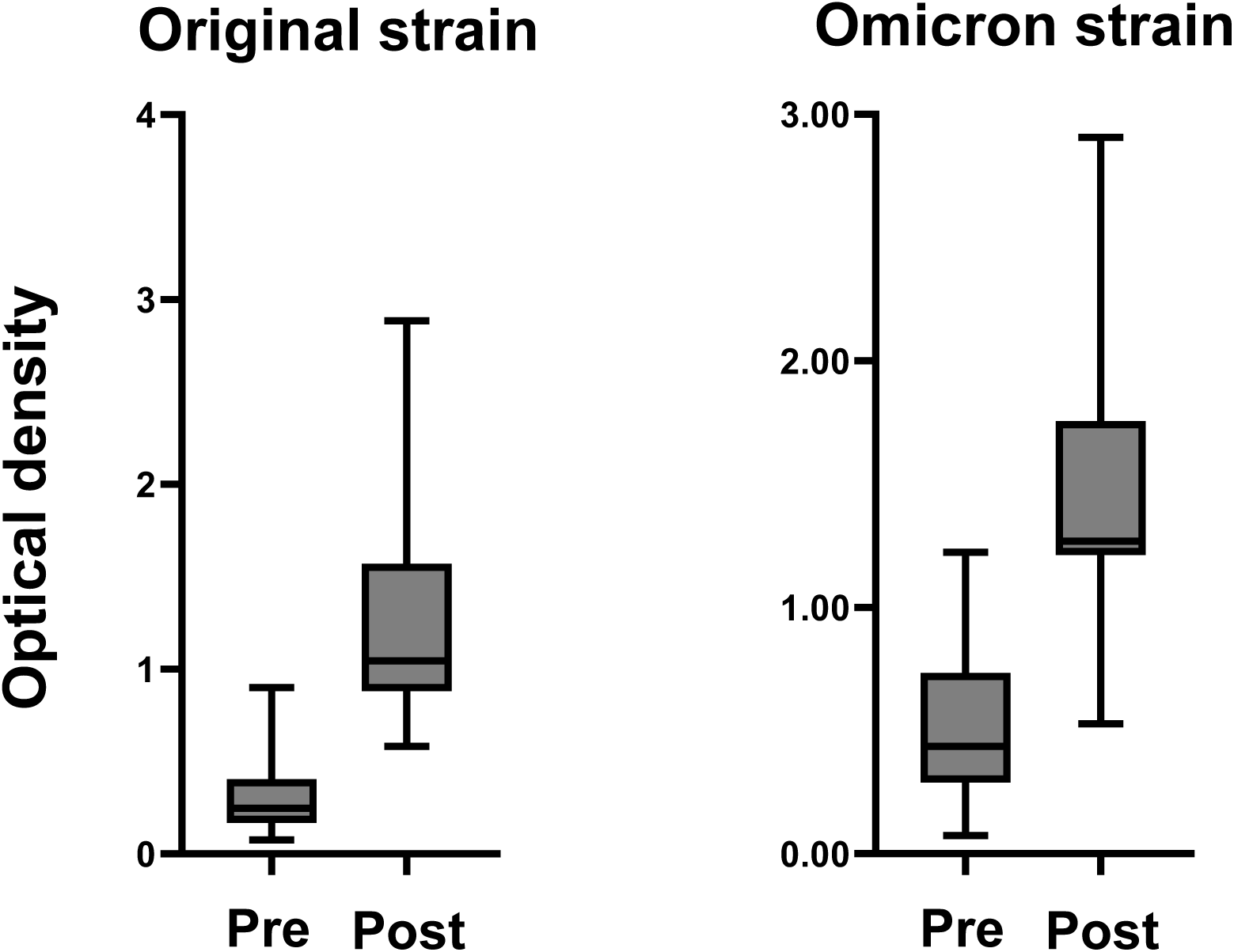
Measurement of anti-spike antibody. Anti-spike antibody of original and Omicron strain titers measured before and after administration of the monovalent Omicron XBB.1.5 mRNA vaccines using fully automated commercial immunoassays in the three participants.

### B-cell receptor repertoire data for quantification of antigen-specific antibody sequence method

To evaluate the immunogenicity of MAFB-7256a vaccination, we analyzed BCR repertoire data pre- and post-vaccination using the database of BCR sequences that bind to SARS-CoV-2 (QASAS method) (Figure 2A).

**Figure 2.**
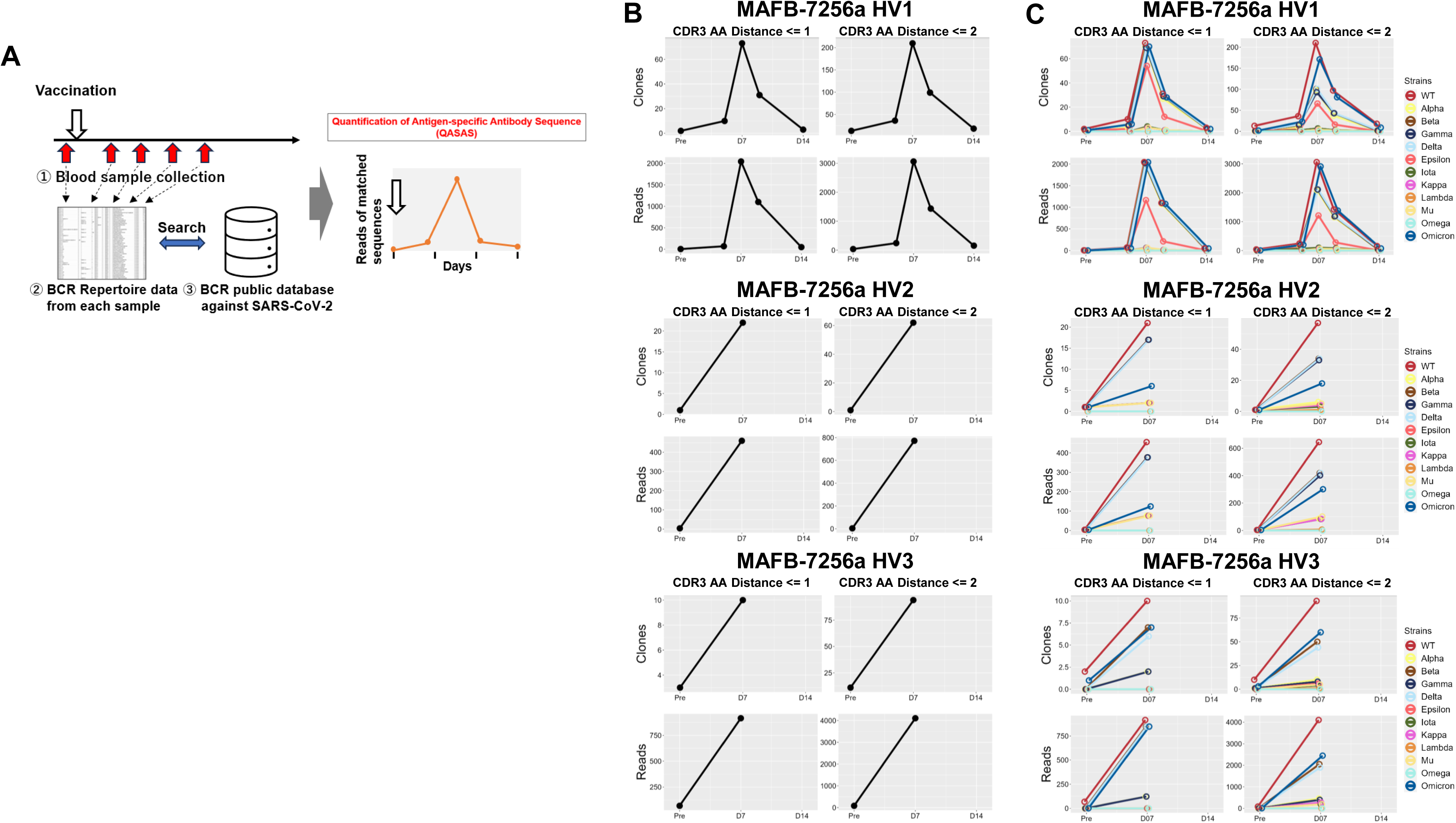
Immunogenicity of MAFB-7256a vaccine evaluated by the Quantification of Antigen-specific Antibody Sequence (QASAS) method. (A) Schematic of the QASAS method. We collected blood samples over time pre- and post-vaccination to analyze the BCR repertoire. We then analyzed the extent to which BCR sequences in the database were contained in the obtained BCR sequence data set, and analyzed the transition of matched BCR sequences. (B) Changes over time in the number (Unique Read, upper) and total reads (Total Read, lower) of SARS-CoV-2-specific sequences following vaccination. Before and after vaccination, SARS-CoV-2-specific sequences were retrieved from the BCR repertoire data. (C) Strains to which the matched sequences bound were analyzed, and the number of matched unique sequences and reads for each strain were counted. Binding specificities for 12 SARS-CoV-2 strains (WT, Alpha, Beta, Gamma, Delta, Epsilon, Iota, Kappa, Lambda, Mu, Omega, and Omicron) in the database were searched by search string. LV, Levenshtein.

Nine blood samples from three participants (MAFB-7256a HV1: pre-, 5, 7, 9, and 14 days after vaccination, MAFB-7256a HV2 and HV3: pre-, and 7 days after vaccination) were analyzed for BCR repertoire. A total of 1,371,046 sequences were obtained, of which 1,341,987 in-frame sequences with V and J genes and CDR3 amino acid assignments were used. We then used the QASAS method to analyze the extent to which SARS-CoV-2-specific sequences were included in the BCR sequence data set obtained from each blood sample of MAFB-7256a-vaccinated donors. Although few matched sequences were detected before vaccination, the number of matched unique sequences and total reads quickly increased 1 week after vaccination (Figure 2B). The QASAS method clearly demonstrated an increase in specific antibodies induced in response to MAFB-7256a.

We then analyzed which strains were bound by each matched sequence, and calculated the number of unique matched sequences along with their total reads for each strain (Figure 2C). Although many of the matched sequences were antibody sequences that bound to WT (original strain), some matched sequences which bound to the Omicron strain were also detected (Figure 2C).

### Epitopes to which each matched BCR/antibody sequence bound

We analyzed which epitopes (Supplementary Table 2) were bound by each matched sequence, and calculated the number of unique matched sequences along with their total reads for each epitope (Figure 3). First, we analyzed matched sequences by the QASAS method in SARS-CoV-2-infected patients. The matched sequences bound to a wide range of epitopes, including the RBD, NTD, and S2 (Figure 3B). We also analyzed matched sequences in individuals vaccinated with BNT162b2, which encodes full spike protein. The matched sequences also bound to a wide range of epitopes, including the RBD, S1 non-RBD, and S2 (Figure 3C). On the other hand, the majority of matched sequences in individuals vaccinated with MAFB-7256a, which encodes an RBD analogue, bound only to the RBD (Figure 3D).

**Figure 3.**
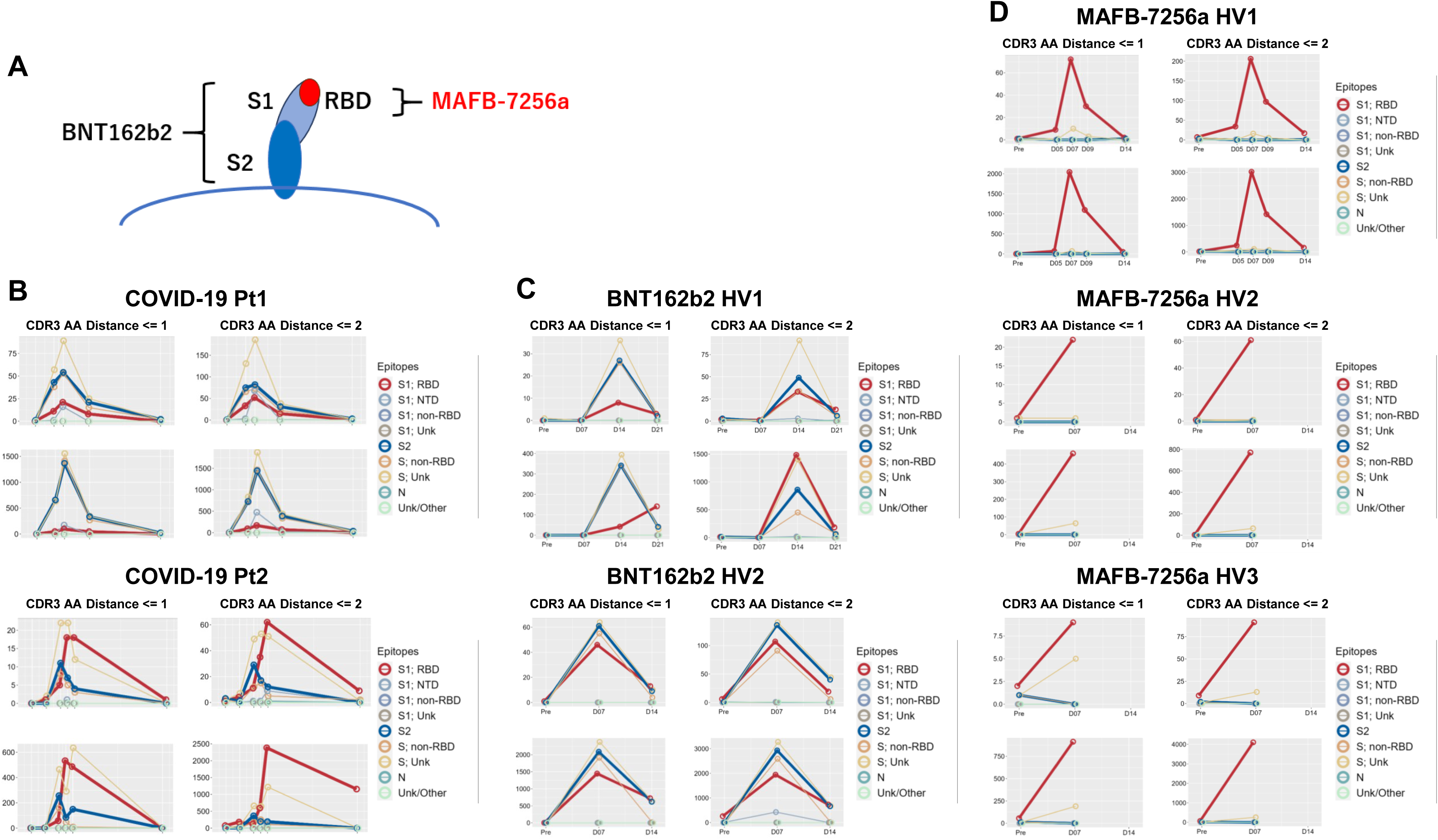
Analysis of epitopes to which the matched BCR/antibody sequences bound. Epitopes to which the matched sequences bound were analyzed, and the number of matched unique sequences and reads for each strain were counted in (A) SARS-CoV-2 infected patients (COVID-19 Pt1 and 2); (B) healthy volunteers vaccinated with BNT162b2, which encodes full spike protein (BNT162b2 HV1 and 2); and (C) healthy volunteers vaccinated with MAFB-7256a, which encodes an RBD analogue (MAFB-7256a HV1, 2 and 3). Binding specificities for 9 SARS-CoV-2 epitopes (S1; RBD, S1; NTD, S1; non-RBD, S1; Unk, S2, S; non-RBD, S; Unk, Nm Unk/Other) in the database were searched by search string. LV, Levenshtein.

We evaluated the immune response, including data from previous studies^8,9^ that used the QASAS method on individuals who had been vaccinated or infected with SARS-CoV-2. Specifically, we compared the binding of matched sequences to the RBD between two groups: nine cases (consisting of three cases of SARS-CoV-2 infection in early 2020 and six cases of individuals who had received vaccines encoding full-length spike protein, including monovalent original mRNA vaccine, bivalent original/Omicron BA.4/5 mRNA vaccine and monovalent Omicron XBB1.5. mRNA vaccine) versus three individuals who had received MAFB-7256a. When we analyzed these cases for sequences showing apparent binding to the RBD or binding to epitopes other than the RBD (Supplementary Table 3), data were not identified to suggest that the matched sequences bound to epitopes other than the RBD (Figure 4).

**Figure 4.**
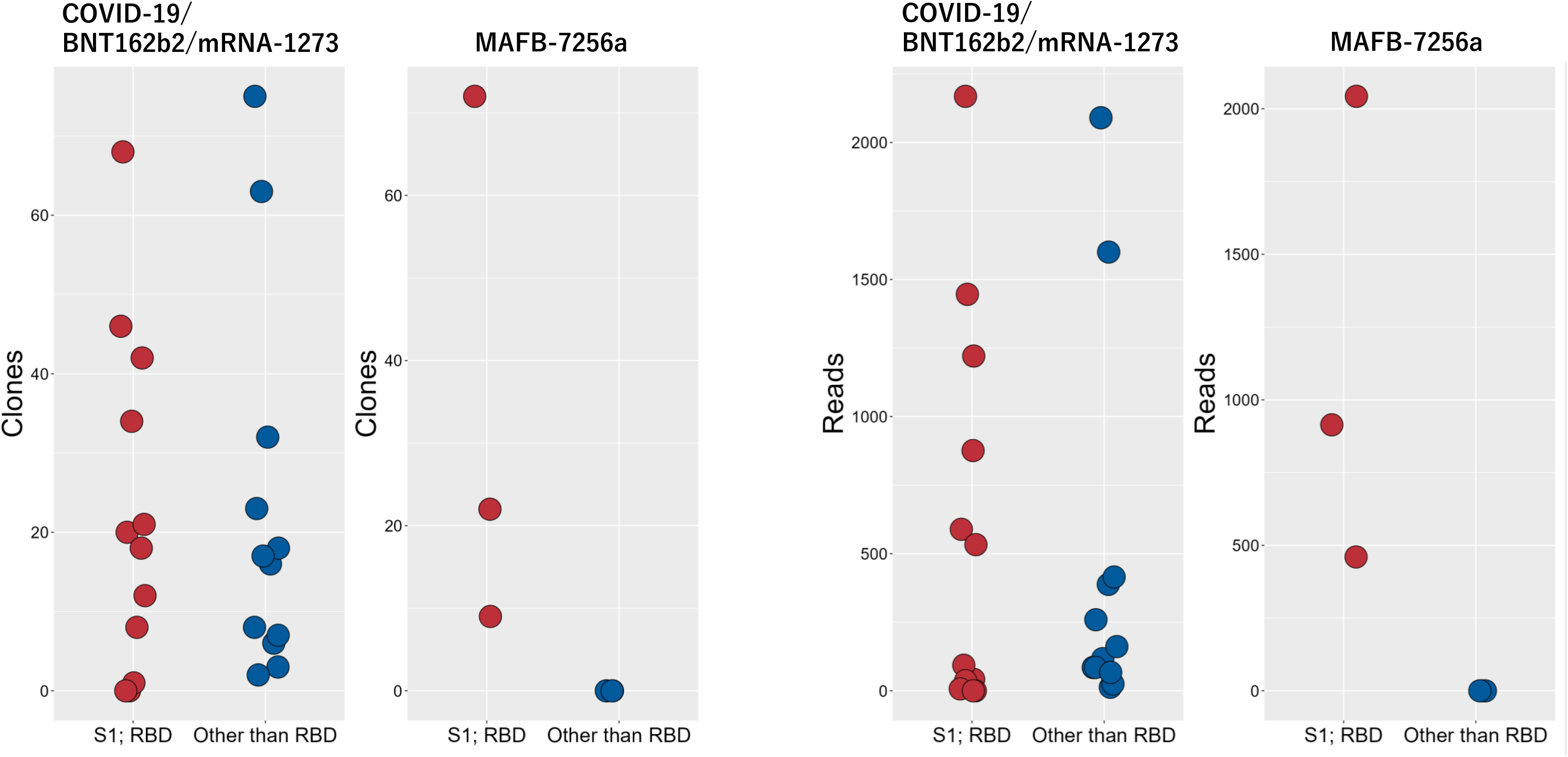
Comparison of MAFB-7256a encoding an RBD analogue with other SARS-CoV-2 antigens (SARS-CoV-2 infection, BNT162b2 and mRNA-1273 encoding full-length spike protein) Three cases of SARS-CoV-2 infection and eleven cases of individuals who received BNT162b2 or mRNA-1273 encoding full-length spike protein were analyzed using the QASAS method. In reads of the matched sequences binding to the RBD and those clearly binding to regions other than the RBD (CDR3 AA Distance ≤ 1), these 14 cases were compared with current MAFB-7256a recipients.

## DISCUSSION

In this study, we assessed the immunogenicity of the monovalent Omicron XBB.1.5 mRNA RBD analogue vaccine MAFB-7256a using the Quantification of Antigen-specific Antibody Sequence (QASAS) method in three healthy volunteers. Our results indicated a rapid increase in matched sequences one week after vaccination, highlighting the vaccine’s ability to induce an immune response. Of note, the majority of matched sequences after vaccination bound to the RBD, and sequences binding to other epitopes were not observed, thereby confirming the targeted action of MAFB-7256a.

The RBD of the S protein is crucial for the virus’s ability to bind to ACE2 receptor, facilitating membrane fusion and cell entry.^6,7^ This makes the RBD a prime target for neutralizing antibodies (nAbs), which are central to the defense against SARS-CoV-2. Previous studies have shown that approximately 90% of nAbs produced by COVID-19 patients target the RBD, emphasizing its importance of targeting the RBD in vaccine development.^13,14^ Our study supports this focus, as MAFB-7256a, which encodes the RBD, effectively stimulated an immune response specifically targeting this critical viral component.

The QASAS method offers advantages over traditional serological tests by providing detailed information on the binding specificity of B-cell receptor (BCR) sequences to various strains and epitopes of SARS-CoV-2. In our study, the QASAS method demonstrated the presence of BCR sequences that bind specifically to the Omicron strain, further validating the effectiveness of MAFB-7256a against this variant. Moreover, CoV-AbDab also contains binding information of BCR/antibody sequences to specific epitopes of SARS-CoV-2. Therefore, we used this analytical method to compare the matched sequences that emerged after SARS-CoV-2 infection or vaccination with vaccines encoding the full-length spike protein and those that emerged after MAFB-7256a vaccination. The results indicated that many of the matched sequences emerging after MAFB-7256a vaccination bound to the RBD, and no matched sequences were observed binding to epitopes other than the RBD. These findings demonstrate the utility of the QASAS method in vaccine evaluation and may support the continued development of RBD-based vaccines for effective SARS-CoV-2 immunization strategies.

In conclusion, we were able to evaluate the immunogenicity of MAFB-7256a using our unique QASAS method, which demonstrated that the vaccine produced expected antibodies capable of binding to the RBD of the Omicron strain.

## Author contributions

Designed research, Y.F., K.Y, G.O., T.M. and H.M.; performed research, Y.F., K.Y, G.O., T.S., S.M., T.Koyama., Y.N., K.K., S.K., N.K. and H.M; contributed vital new analytical tools, T.M., T.S. and T.Kusakabe; analyzed data, Y.F. K.Y, G.O., T.M., T.S. and T.Kusakabe; and wrote the paper, Y.F. K.Y, G.O., T.M. S,M., T.Koyama, Y.N., K.K., S.K, N.K. and H.M.

## Data Availability Statement

All data produced in the present study are available upon reasonable request to the authors.

## Ethics statement

This study was conducted in accordance with the principles of the Declaration of Helsinki.

## Participant consent statement

All participants provided written informed consent for this research.

## Clicical trial registration number

The study protocol was approved by Kobe University Hospital Ethics Committee (No. B2356701).

## Permission to reproduce material from other sources

N/A

---

**Supplementary Table 1.**
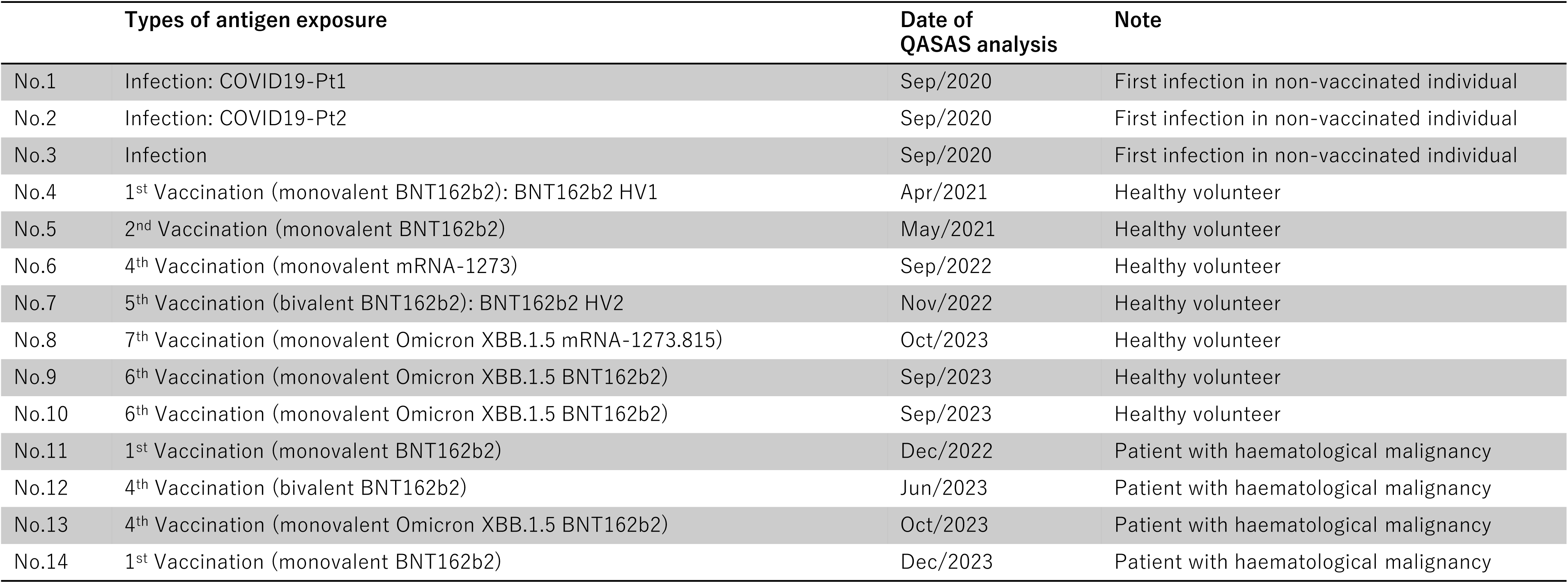
Cohort exposed to SARS-CoV-2 infection, BNT162b2 and mRNA-1273, which encode SARS-CoV-2 full-length spike protein.

**Supplementary Table 2.** Types of epitopes contained in COV-AbDab were classified as shown for adaptation to the QASAS method.

|  |  |
| --- | --- |
| **S1; RBD** | **S; RBD, S; Possibly RBD, S; probably RBD (implied by clustering),** |
| **S1; NTD** | **S; NTD** |
| **S1; non-RBD** | **S; S1 non-RBD, S1; non-RBD** |
| **S1; Unk** | **S;S1** |
| **S2** | **S; S2, S; non-S1, S; S2' Cleavage Site/Fusion Peptide NTD, S; S2 Stem Helix, S; Stem Helix, S; S2 Fusion Peptide** |
| **S; non-RBD** | **S; non-RBD** |
| **S; Unk** | **S; Unk, S; RBD/non-RBD,** |
| **N** | **N, N; NTD,** |
| **Unk/Other** | **Unknown, others** |

**Supplementary Table 3.** Types of epitopes in CoV-AbDab were classified as those showing obvious binding to the RBD or to epitopes other than the RBD.

|  |  |
| --- | --- |
| S1; RBD | S; RBD, S; Possibly RBD, S; probably RBD (implied by clustering), |
| Other than RBD | S; non-RBD, S; S1 non-RBD, S1; non-RBD, S; NTD, S; S2, S; non-S1, S; S2' Cleavage Site/Fusion Peptide NTD, S; S2 Stem Helix, S; Stem Helix, S; S2 Fusion Peptide, N, N; NTD |

